# Is the quality of hospital EHR data sufficient to evidence its ICHOM outcomes performance in heart failure? A pilot evaluation

**DOI:** 10.1101/2021.02.04.21250990

**Authors:** Hannelore Aerts, Dipak Kalra, Carlos Saez, Juan Manuel Ramírez-Anguita, Miguel-Angel Mayer, Juan M. Garcia-Gomez, Marta Durá Hernández, Geert Thienpont, Pascal Coorevits

## Abstract

There is increasing recognition that healthcare providers need to focus attention, and be judged against, the impact they have on the health outcomes experienced by patients. The measurement of health outcomes as a routine part of clinical documentation is probably the only scalable way of collecting outcomes evidence, since secondary data collection is expensive and error prone. However, there is uncertainty about whether routinely collected clinical data within EHR systems includes the data most relevant to measuring and comparing outcomes, and if those items are collected to a good enough data quality to be relied upon for outcomes assessment, since several studies have pointed out significant issues regarding EHR data availability and quality.

In this paper, we first describe a practical approach to data quality assessment of health outcomes, based on a literature review of existing frameworks for quality assessment of health data and multi-stakeholder consultation. Adopting this approach, we perform a pilot study on a subset of 21 International Consortium for Health Outcomes Measurement (ICHOM) outcomes data items from patients with congestive heart failure. To this end, all available registries compatible with the diagnosis of heart failure within the IMASIS-2 data repository connected to the Hospital del Mar network (142,345 visits of 12,503 patients) were extracted and mapped to the ICHOM format. We focus our pilot assessment on five commonly used data quality dimensions: completeness, correctness, consistency, uniqueness and temporal stability.

Overall, this pilot study reveals high scores on the consistency, completeness and uniqueness dimensions. Temporal stability analyses show some changes over time in the reported use of medication to treat heart failure, as well as in the recording of past medical conditions. Finally, investigation of data correctness suggests several issues concerning the proper characterization of missing data values. Many of these issues appear to be introduced while mapping the IMASIS-2 relational database contents to the ICHOM format, as the latter requires a level of detail which is not explicitly available in the coded data of an EHR.

To truly examine to what extent hospitals today are able to routinely collect the evidence of their success in achieving good health outcomes, future research would benefit from performing more extensive data quality assessments, including all data items from the ICHOM heart failure standard set, across multiple hospitals.

## 1. Introduction

Increasing quantities of health data are being collected across care organizations, creating a powerful opportunity to learn from these data how to improve patient care and accelerate research. The earliest call to action and formalized approach for using health data to assess quality of care was probably the Donabedian model of quality ^1^. He categorized the assessment of healthcare quality under structure (how services are organized and resourced), process (how care is delivered and what care activities are undertaken), and outcome (what health impact it has). Over the decades, it has proved much easier to develop and implement audits of structure or process, but formalized assessments of outcome appear to be more challenging because it is harder to define what we mean by outcomes and how best to measure them ^2,3^. A formalized approach to measuring health outcomes was proposed by Porter, within his model of the assessment of “value” in a seminal publication in 2006 ^4^. Within his value equation, outcomes were defined as “the outcomes that matter to patients and the costs to achieve those outcomes” ^4^. This “Value Based Health Care” model has grown into a portfolio of health outcomes standards for measuring value, developed and promoted by the International Consortium for Health Outcomes Measurement (ICHOM) (www.ichom.org). These health outcomes standards, formalized as indicators to be collected, quantified and compared between healthcare providers, have stimulated a global interest in benchmarking and comparing health outcomes ^5^.

All of these models hinge upon the essential ability to measure health, healthcare and its outcomes. Health data is therefore a vital ingredient. To enable accurate measurement, data has to be captured and represented to a high quality. Unreliable data, such as incomplete, incorrect or missing data entries, will inevitably lead to biased analyses, resulting in misdirected efforts to improve quality or false research interpretations. The importance of high-quality data has grown in recognition in recent years. For example for the development of artificial intelligence (AI), the European Commission Ethics guidelines for trustworthy AI state that “The quality of the data sets used is paramount to the performance of AI systems” ^6^.

Yet, several studies have pointed out significant issues regarding availability and quality of EHR data ^7–15^. For example, the “Electronic Health Records for Clinical Research” project, funded by the Innovative Medicine Initiative, clearly demonstrated that many variables, among which even fundamental ones such as patient weight, are frequently not present within EHR systems ^10^. Incorrect or absent recording of patient weights, though, can lead to medication dosage errors. Hirata and colleagues ^16^ examined the frequency and consequences of weight errors that occurred across 79,000 emergency department encounters of children under the age of 5. They revealed that, although weight errors were relatively rare (0.63%), a large proportion of weight errors led to subsequent medication-dosing errors (34%). An earlier study by Selbst and colleagues ^17^ also investigated the consequences of medication errors in a paediatric emergency department. They found that almost half of patients required additional monitoring (30%), examination (6%) or treatment (12%) after medication errors resulting from weight errors. To obtain reliable outcome measures from routinely collected EHR data, Sáez et al. ^15^ have developed a national, standardized and data quality assessed integrated data repository on maternal-child care. During this process, they found that the variability in data quality across hospital sites could lead to unprecise comparison of measurements. Moreover, data quality indices, the efficiency of research processes and the reliability of subsequent results have been found to improve if patient records are assessed for data quality ^9,18,19^. Hence, quality assessment of source health data is crucial to identify and mitigate data quality problems for proper data use and reuse.

In this paper, we first describe our practical approach to quality assessment of health outcomes data. Adopting this methodology, we perform a pilot study on a subset of ICHOM outcomes data collected during routine clinical care of patients with congestive heart failure in a general hospital, given the high prevalence and margin for outcomes improvement in heart failure ^20^. Assessing data quality of outcomes data obtained during routine clinical care is of great interest since currently ICHOM indicators are collected through dedicated data collection into specialist outcome measurement systems, which results in useful data but is not a scalable process.

## 2. Materials and Methods

### 2.1 Data quality assessment

Research into data quality has gained attention since the seminal work by Wang and Strong ^21^, who proposed a comprehensive “fit-for-use” data quality assessment framework using data quality dimensions. Since then, several studies have aimed to define data quality dimensions and methodologies to describe and measure the complex multidimensional aspects of data quality ^19,22–25^. Across studies, little agreement exists about the exact definition and meaning of data quality dimensions. Despite differences in terminology, though, many of the proposed dimensions and solutions aim to address conceptually similar data quality features ^19^.

Following a review of existing literature, the data quality task force of the European Institute for Innovation through Health Data (i∼HD) ^26^ identified nine frameworks for quality assessment of health data ^7,19,24,27–32^. From these frameworks, nine data quality dimensions were selected and prioritized during a series of workshops with clinical care, clinical research and ICT leads from 70 European hospitals. The selected data quality dimensions were deemed most important to assess the quality of health data if this data is to be useful for patient care, for organizational learning (quality improvement, such as the assessment of health outcomes) and research (big data research and case finding for clinical trials recruitment). Table 1 provides an overview of the selected data quality dimensions, together with their original terminology.

**Table 1.**
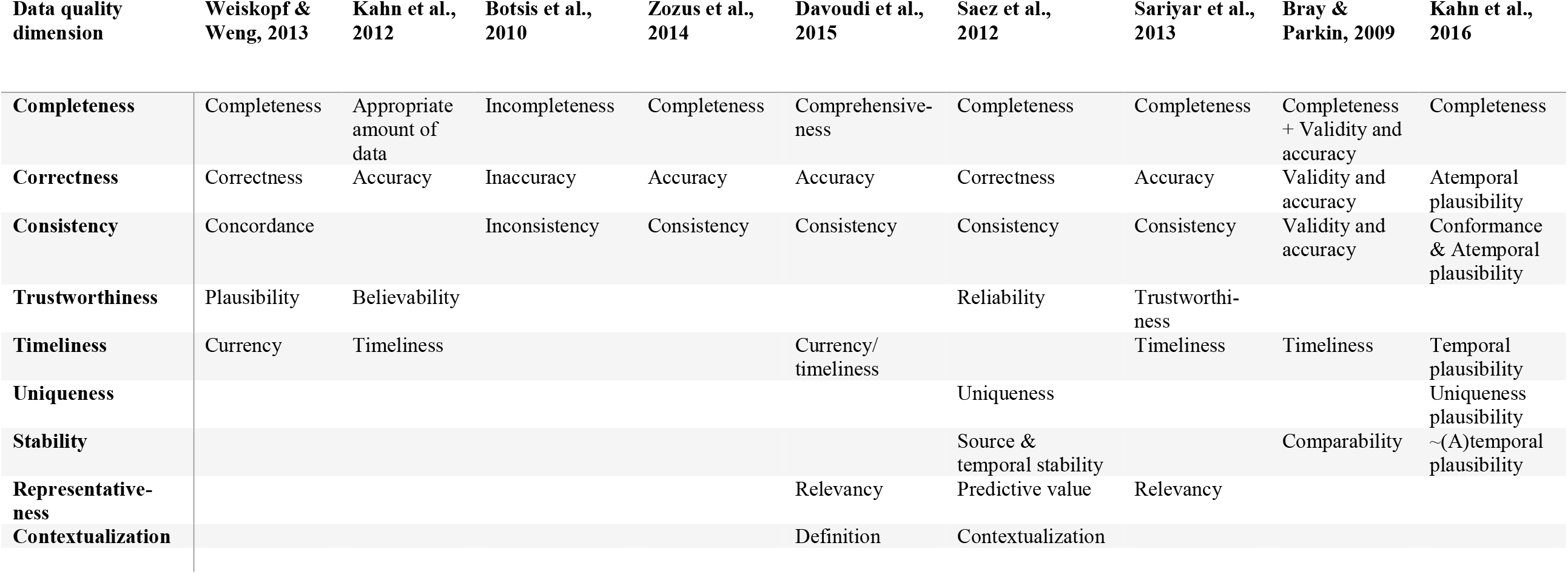
Mapping of data quality dimensions.

| **Data quality dimension** | **Weiskopf & Weng, 2013** | **Kahn et al., 2012** | **Botsis et al., 2010** | **Zozus et al., 2014** | **Davoudi et al., 2015** | **Saez et al., 2012** | **Sariyar et al., 2013** | **Bray & Parkin, 2009** | **Kahn et al., 2016** |
| --- | --- | --- | --- | --- | --- | --- | --- | --- | --- |
| **Completeness** | Completeness | Appropriate amount of data | Incompleteness | Completeness | Comprehensive-ness | Completeness | Completeness | Completeness + Validity and accuracy | Completeness |
| **Correctness** | Correctness | Accuracy | Inaccuracy | Accuracy | Accuracy | Correctness | Accuracy | Validity and accuracy | Atemporal plausibility |
| **Consistency** | Concordance |  | Inconsistency | Consistency | Consistency | Consistency | Consistency | Validity and accuracy | Conformance & Atemporal plausibility |
| **Trustworthiness** | Plausibility | Believability |  |  |  | Reliability | Trustworthi-ness |  |  |
| **Timeliness** | Currency | Timeliness |  |  | Currency/timeliness |  | Timeliness | Timeliness | Temporal plausibility |
| **Uniqueness** |  |  |  |  |  | Uniqueness |  |  | Uniqueness plausibility |
| **Stability** |  |  |  |  |  | Source & temporal stability |  | Comparability | ~(A)temporal plausibility |
| **Representative-ness** |  |  |  |  | Relevancy | Predictive value | Relevancy |  |  |
| **Contextualization** |  |  |  |  | Definition | Contextualization |  |  |  |

The completeness, consistency, correctness and uniqueness dimensions are commonly used in the data quality literature ^19,25^. Although sometimes the first three can overlap on their definitions, or be contained within each other, we prefer making them orthogonal. For instance, a patient observation is incomplete if it is not registered, inconsistent if it does not comply with formatting requirements, or incorrect if it is unlikely to be true for a specific patient. For example, multiple normal kidney blood test results in a patient on dialysis would be consistent, though incorrect. Uniqueness, in turn, assesses whether duplications are present among patient records, for example as a result of an incomplete merging of patient records between hospital departments.

Further, stability relates to the probabilistic concordance of data among different data sources such as hospitals, physicians or devices, or over time ^33^. For example, variability among centers has been found in liver offer acceptance rates for pediatric patients, that cannot be explained by donor and recipient factors ^34^. In some cases, standardization of procedures and analyses can reduce levels of variability. However, sometimes differences among centers persist even when using standard procedures, for instance between diffusion tensor magnetic resonance imaging findings obtained at different acquisition centers using a standard protocol ^35^. Likewise, when data is collected over time, temporal changes can occur due to several reasons, including changes in clinical practice or coding scheme used in the EHR ^36^.

Next, timeliness describes how promptly information is processed, or how up-to-date recorded information is; for instance, to evaluate whether a current medication list within an EHR system is up to date or if there is a delay in updating this from a pharmacy sub-system. Trustworthiness relates to the availability of registry governance metadata and the data owner’s reputation. For example, it must be possible for someone accessing a health data item or clinical document to confidently know when and where it was captured, by whom, and if it has been modified since the original entry. Further, contextualization relates to whether the data are annotated with their acquisition context, which can be crucial for correct interpretation of the results. For instance, whether blood glucose laboratory results were obtained while the patient was fasting or not. Finally, representativeness captures whether a dataset is representative for the population it is supposed to be drawn from, in order to allow valid inference.

### 2.2 Pilot assessment

#### 2.2.1 Dataset

##### Data source

For this pilot assessment, we used data from the Parc Salut Mar Barcelona; a complete healthcare services organization with its information system database (IMASIS) as EHR. IMASIS includes and shares clinical information of two general hospitals, one mental health care center, one social-healthcare center and five emergency rooms in the Barcelona city area (Spain). IMASIS contains clinical information from approximately 1.5 million patients who have used the services of this healthcare system since 1989, across different settings such as admissions, outpatient consultations, emergency room visits and major ambulatory surgery appointments. IMASIS-2 is the anonymized relational database of IMASIS, that was created during the European Medical Information Framework (EMIF) project ^37^, being the data source used for research purposes. It contains structured data related to diagnosis, procedures, drug administration and laboratory tests, and clinical annotations in a free text format. Since natural language processing falls beyond the scope of this project, we only used structured data. The study protocol was approved by the Ethics Committee of Parc Salut Mar (num. 2016/6935/I), under the research activities related to ischemic heart disease, carried out during the EMIF project funded by the Innovative Medicines Initiative.

##### Patients

As a case study, data of patients diagnosed with congestive heart failure (CHF) were used. Heart failure is a chronic condition, severely impacting people’s quality of life. With a prevalence of over 23 million worldwide, it poses a significant public health problem ^38^. Collecting meaningful data on the health status of heart failure patients is therefore an important step to ensure better quality care, and as a result better quality of life, for these patients.

All patients who attended the hospital at least once between January 1^st^, 2006 and November 7^th^, 2017 and who had at least one diagnosis entry of CHF were extracted from the IMASIS-2 database. Specifically, the selection of patients was based on the following diagnosis codes of the International Classification of Diseases ninth edition (ICD-9): 428, 428.0, 428.1, 428.2, 428.20, 428.21, 428.22, 428.23, 428.3, 428.30, 428.31, 428.32, 428.33, 428.4, 428.40, 428.41, 428.42, 428.43, 428.9. In total, the dataset includes 142,345 patient visit records, describing the medical history of 12,503 different patients.

##### Variables

The ICHOM heart failure outcomes standard set ^39^ was chosen as the most appropriate source of outcome indicators to target. Out of the total of 72 ICHOM data items, a subset of 21 variables was selected as being most likely to be routinely collected within the hospital in patients suffering from CHF, and to be indicative of the overall quality of data collected on this type of patients. In addition, a visit identifier was included, to distinguish different patient visit records. An overview of all variables included in the pilot assessment can be found in Table 2. In addition, Supplementary Tables 1 and 2 show the ICD-9 codes used to identify baseline health status variables, and the Anatomical Therapeutic Chemical classification system (ATC/DDD) codes of the World Health Organization ^40^ to retrieve patients’ medication usage, respectively.

**Table 2.** Overview of ICHOM variables used in pilot assessment.

| Item | Definition | Response options |
| --- | --- | --- |
| **Identifiers** |  |  |
| Patient ID | Patient's medical record number | According to institution |
| Visit ID | Unique visit record identifier | [Not included in ICHOM standard set] |
| **Demographic factors** |  |  |
| Age | Date of birth | DD/MM/YYYY |
| Sex | Sex at birth | 1 = Male<br>2 = Female |
| **Baseline health status** |  |  |
| Atrial fibrillation | Ever diagnosed with atrial fibrillation | 0 = No<br>1 = Yes<br>999 = Unknown |
| Prior MI | Ever diagnosed with myocardial infarction | 0 = No<br>1 = Yes<br>999 = Unknown |
| Hypertension | History of hypertension | 0 = No<br>1 = Yes<br>999 = Unknown |
| Diabetes mellitus | Ever diagnosed with diabetes mellitus | 0 = No<br>1 = Yes<br>999 = Unknown |
| Echocardiogram performed | Echocardiogram performed to assess ejection fraction | 0 = No<br>1 = Yes<br>999 = Unknown |
| Height | Height | Numeric value of height in metric system |
| Weight | Weight | Numeric value of weight in metric system |
| Alcohol use | Consumption of >1 alcoholic drink a day | 0 = No<br>1 = Yes<br>999 = Unknown |
| Smoking status | Current smoking status | 0 = No<br>1 = Yes<br>999 = Unknown |
| **Treatment variables** |  |  |
| Beta Blocker | Beta blockers currently prescribed for heart failure | 0 = No<br>1 = Yes<br>999 = Unknown |
| Calcium Channel Blocker | Calcium channel blockers currently prescribed for heart failure | 0 = No<br>1 = Yes<br>999 = Unknown |
| Digoxin | Digoxin currently prescribed for heart failure | 0 = No<br>1 = Yes<br>999 = Unknown |
| Diuretics | Diuretics currently prescribed for heart failure | 0 = No<br>1 = Yes<br>999 = Unknown |

| Burden of care |  |  |
| --- | --- | --- |
| Date of arrival | Date of admittance | DD/MM/YYYY |
| Date of discharge | Date of discharge | DD/MM/YYYY |
| Hospital admissions | Number of hospitalizations in last 12 months due to heart failure | Numerical value or 999 = Unknown |
| Hospital appointments | Number of hospital appointments in last 12 months due to heart failure | Numerical value or 999 = Unknown |
| Mortality |  |  |
| Date of death | Date patient was declared dead | DD/MM/YYYY or 999 = Unknown |

Figure 1 provides a graphical summary of the different steps that were performed in order to obtain our study dataset.

**Figure 1.**
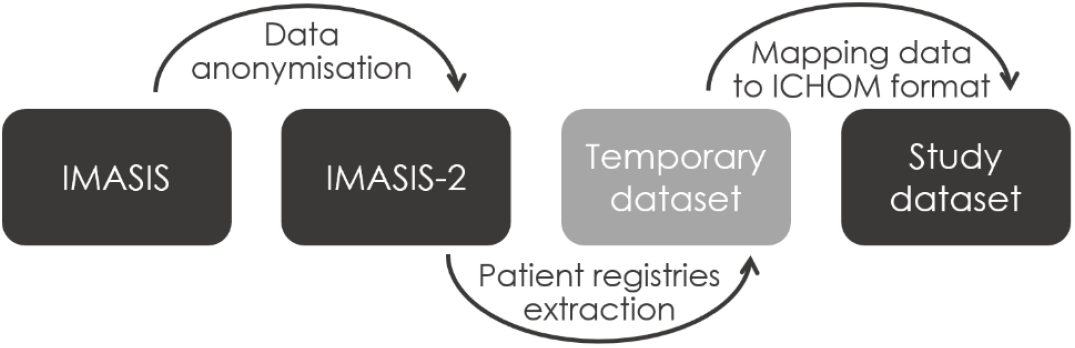
Overview of procedure to generate study dataset. From the original EHR system (IMASIS), patient data are anonymized during transfer to the IMASIS-2 database. For our study, we then extracted data providing information about 21 ICHOM items from heart failure patient registries from the IMASIS-2 database, after which all data items were mapped to their corresponding ICHOM response format.

#### 2.2.2 Data quality dimensions

To evaluate the quality of heart failure patient data collected during routine clinical care, a subset of five data quality dimensions was selected by the hospital: completeness, correctness, consistency, uniqueness and stability. These dimensions are most commonly used in the data quality literature and were deemed most interesting to assess given the nature of the data.

First, we investigated the frequency with which partially duplicated patient records occur. Second, we assessed consistency by data type, range and basic multivariate rules (for example: patients’ arrival date should be before or equal to their date of discharge) ^15^. Next, we investigated data completeness by computing the proportion of complete fields per variable. Further, we qualitatively evaluated temporal stability of recorded past medical conditions and usage of different types of medications by computing their respective relative frequencies on a monthly basis to visualize trends over time. Finally, we inferred data correctness, either by combining information across variables or by investigating data from the same patient over time.

#### 2.2.3 Tools

We conducted data quality assessment using R, version 3.6.1 ^41^. For the temporal stability analyses, we used the EHRtemporalVariability R package ^42^.

## 3. Results

### 3.1 Uniqueness

Out of a total of 142,345 patient visits records, 1.2% have identical visit identifiers even though values on one or more data items have different inputs. In turn, 2.8% of all patient visit records have identical data while the visit identifier differed. This amounts to a total score of 96% for uniqueness.

### 3.2 Consistency

Consistency by type and by multivariate rules both yield a score of 100%; all values are in the right format and no errors in relationships between dates are found. As a third consistency check, we examine whether numerical and date values fall within pre-specified ranges, and whether categorical variables have values that comply with pre-defined response options. An average score of 91.21% is obtained for consistency by range, resulting from errors in three variables. In particular, 85% of values for height and weight are “0”. Since weight and height values of zero do not have a physical meaning, we hypothesise these data points are missing data values. This was confirmed by data management staff at the Hospital del Mar, clarifying that zero entries are not even permitted in the structured data fields of height and weight. Rather, these zero values were introduced during data extraction from the IMASIS-2 database to indicate missingness, since only numeric values are accepted for height and weight according to the ICHOM Heart Failure data dictionary (summarised in Table 2). In addition, a small number of out-of-range data points are identified for height (54) and weight (20). Further, 16 visit records have arrival dates before January 1, 2006. In sum, this yields a consistency score of 97.07%.

### 3.3 Completeness

Assessing completeness of the dataset by column reveals that all included variables are entirely documented, except for date of death which is only recorded in 37.14% of all patient visits. Data management staff at the Hospital del Mar confirmed that this incompleteness is valid, since date of death is only provided in case the patient died during the visit. Excluding the latter valid incompleteness result, an average score of 100% is obtained for completeness.

#### 3.4 Temporal stability

Two categories of data items are assessed for temporal variability: medication usage and past medical conditions. As illustrated in Figure 2, results show a gradual increase over time in the recorded usage of different types of medication to treat heart failure, especially of beta blockers and diuretics. Further, we find an abrupt change in documentation pattern of past medical conditions in 2011, with drastically reduced frequencies of reported past medical conditions (Figure 3). Of note, only a small number of patient visit records (<10) is available each month in the first half of 2016, explaining the absent or divergent results.

**Figure 2.**
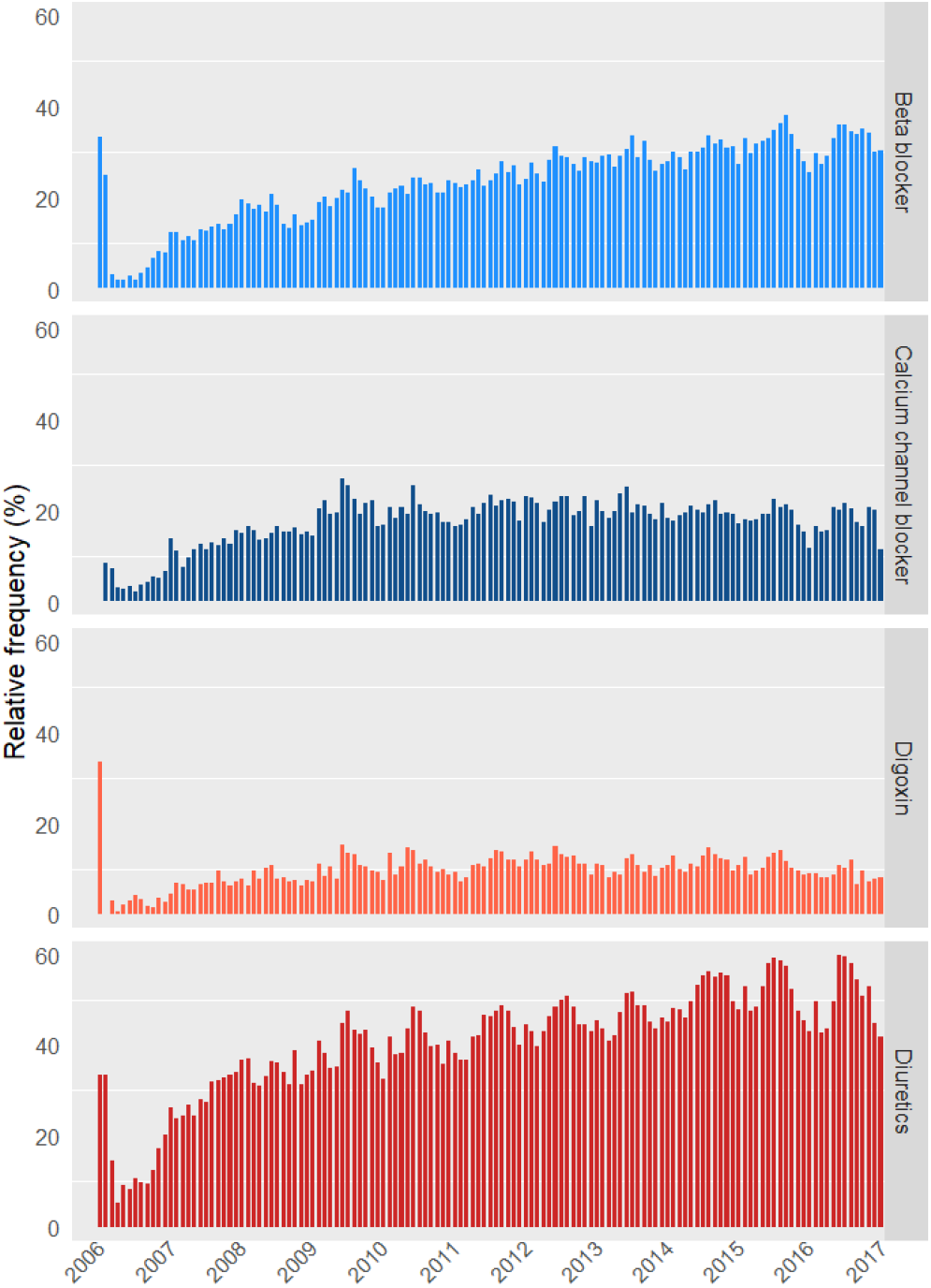
Relative frequencies of recorded drug usage per month, plotted over time.

**Figure 3.**
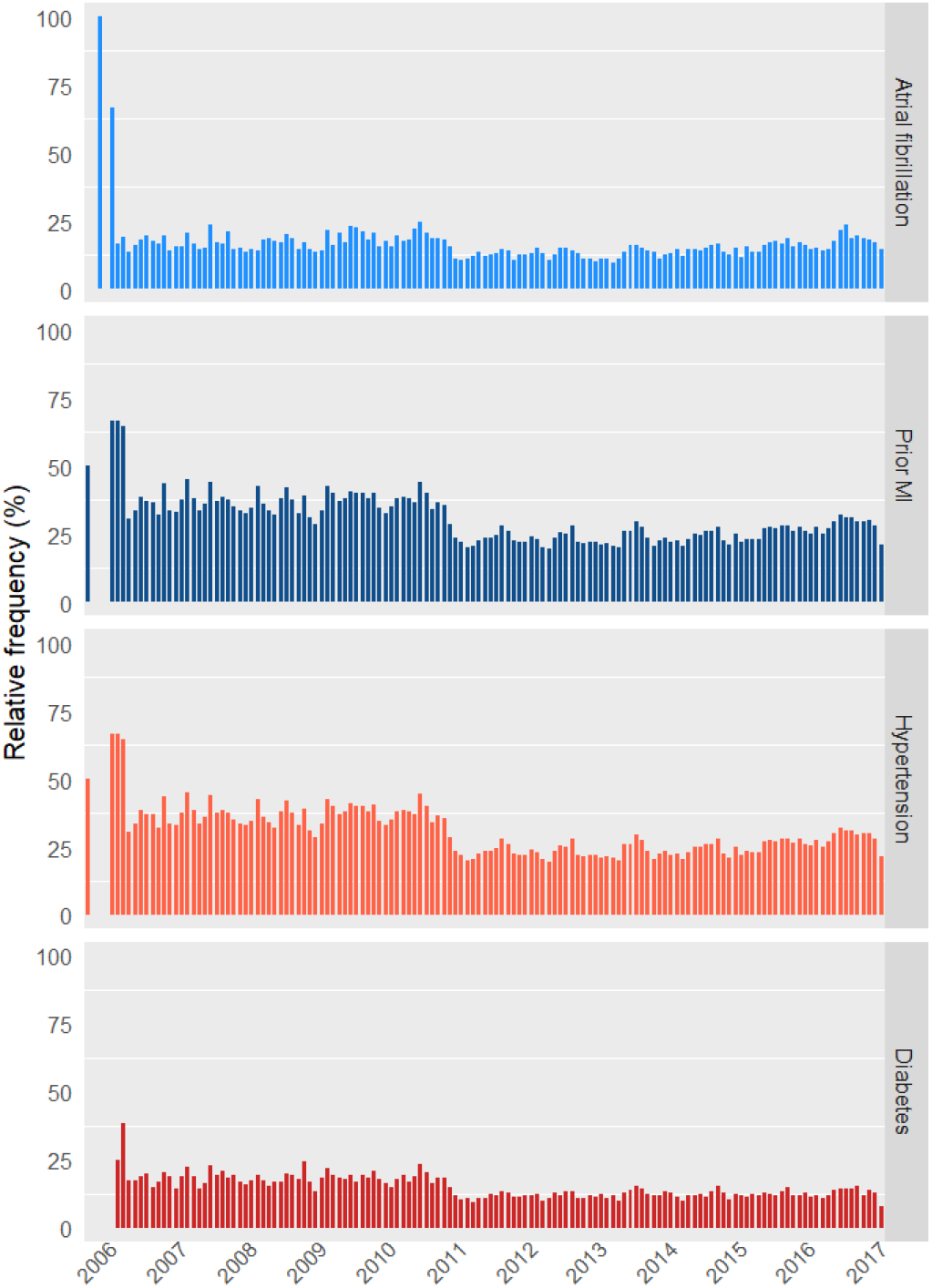
Relative frequencies of recorded past medical conditions per month, plotted over time.

### 3.5 Correctness

After performing some basic descriptive analyses, results of which are summarized in Supplementary Table 3, two sets of variables are subjected to closer inspection. Firstly, correctness of height and weight values is evaluated based on their bivariate distribution as shown in Figure 4. All data points that fall below the main diagonal, implying that patients’ weight (in kg) is larger than their height (in cm), are very unlikely to be true. A subset of these data errors, highlighted by the red circle, are hypothesized to result from value inversion between height and weight recordings. To formally assess implausible height and weight values, we compute patients’ body mass index (BMI). Results show that 16 patients have a suspiciously low BMI (< 10), and 180 patients have an implausibly high BMI (> 70). Hence, a total of 196 probable errors are identified, corresponding to 0.13% of all patient visit records.

**Figure 4.**
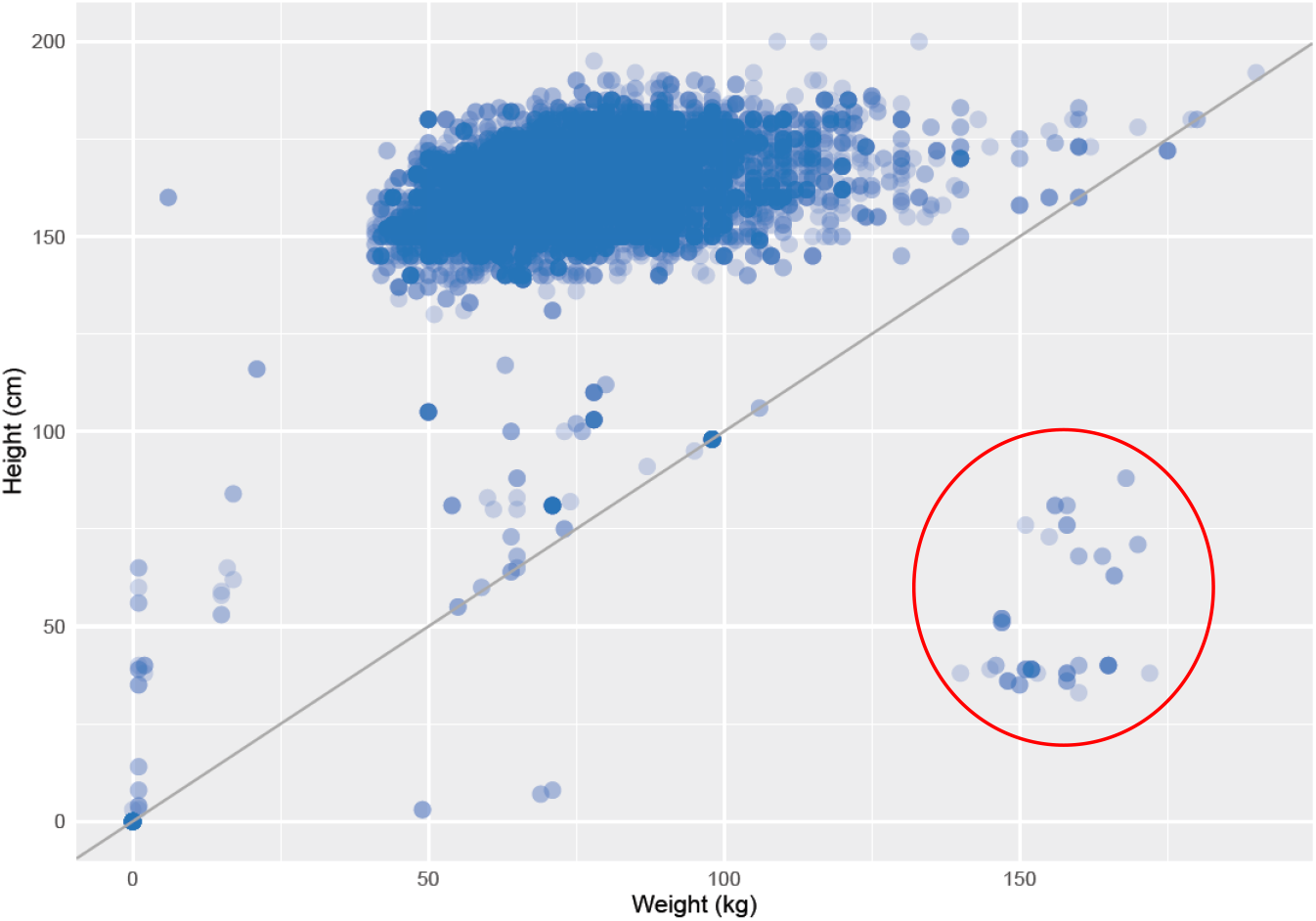
Bivariate distribution of height and weight values, with red circle highlighting data points where height and weight values are hypothesized to have been inverted.

Further, we investigate the temporal order of past medical conditions, assuming that once a hospital visit record indicates that a patient has a history of atrial fibrillation, hypertension, diabetes or myocardial infarction, the history of this diagnosis/event should be mentioned in all subsequent visit records. Deviations from this temporal order (i.e., “history” followed by “no history”) hence point to data errors in the extracted dataset. Results show a substantial amount of deviations. Specifically, 6.33% of all patient visit records mention that the patient does not have a history of atrial fibrillation, while earlier records indicate the patient has been diagnosed with atrial fibrillation before. Similarly for history of hypertension, diabetes mellitus and myocardial infarction, error rates of 12.11%, 6.12% and 12.11% are obtained, respectively. Data management staff at the Hospital del Mar clarified that many of these issues appear to be introduced while mapping the IMASIS-2 relational database contents to the ICHOM format, as the latter requires a level of detail which is not explicitly available in the coded data of an EHR. In particular, diagnosis/events already recorded in a previous visit and not mentioned in a subsequent visit are not consistently recorded in EHR systems during routine clinical care, in contrast to data collected for research purposes. It is therefore practically impossible to distinguish true negative from missing data when extracting data from the EHR. As a result, a substantial proportion of patient history data items that were negative in the dataset actually represent missing data values.

Taken together, this amounts to a total score of 93.84% for correctness.

## 4. Discussion

### 4.1 Data quality assessment results and suggestions for improvement

Overall, this pilot assessment reveals high scores on each of the dimensions used to investigate the quality of heart failure patients’ data. Nevertheless, several data quality issues are identified, based on which we propose a set of improvement strategies.

First, results of our data quality assessment show that a substantial amount of negative values in the dataset – indicating the absence of a particular data item – actually represent missing data. As a consequence, some variable distributions seem to be biased. For example, according to the data, only a minority of patients currently smoke or have a past medical condition such as hypertension (see Supplementary Table 3), which is rather implausible for a population of heart failure patients. This is an intrinsic issue associated to structured data sources in the framework of EHR databases. That is, when a code is not found in the EHR, it is practically impossible to distinguish whether the code is negative (i.e., examination has confirmed the absence of a particular condition) or missing (i.e., no examination has taken place, or examination confirmed the presence of a particular condition but is not recorded in structured format) for a given patient. We are aware that good clinical practice does not mandate the measurement of every data item at each patient visit (e.g., disease history), since these items usually are present as additional information in a typical EHR environment. Nevertheless, this differs fundamentally from data collection practices in the context of research activities such as outcomes assessment, for which the ICHOM standard set was originally developed. When performing analytical and research activities, it would therefore be very useful to introduce mechanisms or tools that allow to differentiate data missingness from true negatives, and to determine the duration of each condition and disease, regardless of whether they are mentioned in each visit.

Moreover, many data items are often recorded in free text rather than structured data fields, making it difficult to extract this information for research and analysis purposes. We therefore advise to maximally include data items in form format or specific fields or sections in the EHR. In addition, when using form formats, we recommend the use of alarms for avoiding missing values, as well as for inputting out of range data. Alternatively, natural language processing techniques applied to free text clinical annotations fields can be used to enrich structured sources.

Further, uniqueness analyses reveal some partially duplicated patient visit records. First, duplications in visit identifiers are found while clinical data show different inputs. Data management staff at the Hospital del Mar clarified that this happened whenever different height and weight measurements were registered during a single visit. If a slight difference between values is observed, partial row duplicates are generated when merging data in the final dataset. Secondly, duplicated rows with different visit identifiers have arisen as a consequence of the particular data organisation in IMASIS-2, where some clinical data are ambiguously connected to visit IDs via date matching. As a result, all clinical data collected during different patient visits on the same day are connected to two different visit IDs. To reduce future data quality issues of this kind, we suggest a data reorganization including a two-level visit structure. First, a more general level describes a period in which one or different visits occur, and is connected to clinical data obtained within this period. Second, a more specific level then describes every distinct visit together with corresponding diagnosis and procedures information obtained at the particular visit. This two-level visit organization would contribute to the elimination of partial replicates, thus positively impacting the uniqueness aspect of data quality. This strategy has been previously adopted by the OMOP CDM standard ^43^ with the aim of easing mappings from ambiguous visit- connected schemas.

Finally, we recommend to prepare carefully for the potential impact of changes or upgrades in EHR (sub)system and diagnostic coding practices. Temporal stability analyses reveal an abrupt change in documentation pattern of past medical conditions in 2011, with drastically reduced frequencies of reported past medical conditions (section 3.4). After consultation with data management staff at the Hospital del Mar, it appears this can be explained by the introduction of a new automated coding system in the emergency department EHR systemt. Although we assume this evolution in the recording of past medical conditions had a positive impact on direct patient care, decision support and alert algorithms can be impacted by changes in diagnostic coding practice and should therefore be taken into account. In addition, these changes will affect the reuse of data for research and quality monitoring such as outcomes tracking. In this sense, quality assessment is an essential tool to detect effects of changes in EHR systems introduced over time, which would contribute to a better understanding of the updates in the content and structure of these types of databases.

### 4.2 Limitations and future directions

In interpreting the results of this study, some important limitations should be taken into consideration. First of all, only a subset of ICHOM outcome variables was assessed for their data quality, as the hospital determined these were most likely to be routinely collected within their EHR in patients suffering from CHF. Whether data quality results from this pilot assessment are generalizable to the complete ICHOM standard set has yet to be investigated. Similarly, we selected five out of nine available data quality dimensions, as these were thought to be most relevant given the nature of the data. Further, data quality assessment was performed on a data extract from the IMASIS-2 dataset, after mapping the data items to the ICHOM outcomes format, which might have introduced additional errors. We therefore recommend future studies to examine data quality on the EHR variables directly, in the hospital’s own response format, or to perform an additional data quality assessment of the mapping procedure.

In sum, future research would benefit from performing more thorough data quality assessments, across multiple hospitals, to truly examine to what extent hospitals today are able to routinely collect the evidence of their success in achieving good health outcomes. The European Federation of Pharmaceutical Industries and Associations (EFPIA) is currently leading such a project together with i∼HD. In particular, the goal of this project is to assess the availability and quality of routinely collected patient data to underpin a future scale up of value-based care models in which ICHOM outcomes indicators serve as the measures of value delivered by healthcare provider organizations. For this project, data of patients with heart failure are also being examined, now using the complete set of ICHOM outcomes indicators and performing assessments across ten European hospitals. The promotion of data quality is essential to advance learning health systems, patient empowerment and clinical research, and the results of this larger project will provide interesting insights on the generalizability of the current pilot project’s findings.

## Data Availability

Due to the retrospective nature of this research, patients of this study did not agree for their data to be shared publicly, so supporting data is not available.

## Authors’ contributions

HA performed data quality analyses, interpreted the results and wrote the manuscript. CS and MDH performed data quality analyses. Baseline data quality assessment scripts in R were provided by CS, MDH and JMGG. All authors interpreted data quality analyses results, contributed to the writing of the manuscript, performed critical revisions of the manuscript and approved the final version for publication. MAM and JMRA selected the variables to be included in the analysis, and provided the data for analysis.

