## Supplementary Table 1 for "Is the quality of hospital EHR data sufficient to evidence its ICHOM outcomes performance in heart failure? A pilot evaluation"

**Supplementary Table 1**. ICD-9 classification codes used for the evaluation of baseline health status variables.

| Variable | ICD-9 classification codes |
|---|---|
| **Hypertension** | 401, 401.1, 401.0, 404.1, 404.10, 404.11, 404.12, 404.13, 402.1, 403.1, 405.1, 403, 403.0, 403.00, 403.01, 401, 401.9, 997.91, 404, 404.0, 404.00, 404.01, 404.02, 404.03, 404.9, 404.93, 404.92, 402, 402.9, 403.1, 403.10, 403.11, 404.9, 404.90, 404.91, 404.92, 404.93, 403.9, 403.90, 403.91, 362.11, 402.0, 402.00, 402.01, 402.10, 402.11, 402.90, 402.91, 405, 405.0, 405.01, 405.09, 405.11, 405.19, 405.99, 405.9, 405.91, 405.91 |
| **Diabetes mellitus** | 250, 250.0, 250.00, 250.01, 250.02, 250.03, 250.1, 250.10, 250.11, 250.1, 250.13, 250.2, 250.20, 250.21, 250.22, 250.23, 250.3, 250.30, 250.31, 250.32, 250.33, 250.4, 250.42, 250.41, 250.43, 250.5, 250.50, 250.52, 250.51, 250.53, 250.6, 250.60, 250.62, 250.61, 250.63, 250.7, 250.70, 250.72, 250.71, 250.73, 250.8, 250.80, 250.82, 250.81, 250.83, 250.9, 250.90, 250.92, 250.91, 250.93, 357.2, 362.0, 362.01, 362.02, 362.03, 362.04, 362.05, 362.06, 362.07, 366.41 |
| **Atrial fibrillation** | 427, 427.3, 427.31, 427.32, 427.9 |
| **Smoking status** | 305.1, V15.82, 649.0, 989.84 |
| **Alcohol use** | 303.9, 303.90, 303.91, 303.92, 303.93, 425.5, 305.0, 305.00, 305.01, 305.02, 305.03, 303.0, 303.00, 303.01, 303.02, 303.03 |
| **Echocardiogram performed** | 88.72, 37.28, 00.24 |
