## Supplementary Table 2 for "Is the quality of hospital EHR data sufficient to evidence its ICHOM outcomes performance in heart failure? A pilot evaluation"

**Supplementary Table 2.** Anatomical Therapeutic Chemical classification system (ATC/DDD) codes of the World Health Organization used to retrieve patients' medication usage.

| Drugs | ATC/DDD classification codes |
|---|---|
| **Beta blocker** | C07AB07, C07AB03, C07AG02, C07AB02, C07AB12, C07AA05, C07AG01, C07AA12, C07AB08, C07AA07, C07AB09 |
| **Calcium channel blocker** | C08DB01, C08CA11, C08DA01, C08CA01, C08CA16, C08CA09, C08CA08, C08CA04, C08CA05, C08CA02, C08CA07, C08CA13, C08CA06, C08CA12 |
| **Digoxin** | C01AA05, C01AA08 |
| **Diuretics** | C03EA04, C03X, C03DA04, C03BA04, C03AA03, C03BX, C03CA04, C03CA01, C03BA11, C03EB01, C03CA02, C03DA01, C03CA03, C03BA10, C03EA01, C03AX91, C03EA06, C03XA, C03DB, C03XA01 |
