## Supplementary Table 3 for "Is the quality of hospital EHR data sufficient to evidence its ICHOM outcomes performance in heart failure? A pilot evaluation"

**Supplementary Table 3.** Results of descriptive analyses.

| Item | Descriptive statistics |
|---|---|
| **Numerical variables** | Median (IQR) |
| Age at admission (years) | 78 (68-84) |
| Age at death (years) | 82 (74-87) |
| Height (cm)* | 163 (155-170) |
| Weight (kg) * | 74 (65-85) |
| Hospital admissions (number) | 0 (0-0) |
| Hospital appointments (number) | 0 (0-0) |
| **Date variables** | Median (range) |
| Date of arrival | 03/03/2013 (23/03/1990-24/07/2017) |
| Date of discharge | 20/03/2013 (03/10/2006-27/09/2017) |
| **Categorical variables** | Proportions |
| Sex | Male (53%); Female (47%) |
| Atrial fibrillation | Yes (15%); No (85%); Unknown (0%) |
| Prior MI | Yes (28%); No (72%); Unknown (0%) |
| Hypertension | Yes (28%); No (72%); Unknown (0%) |
| Diabetes mellitus | Yes (14%); No (86%); Unknown (0%) |
| Echocardiogram performed | Yes (3%); No (97%); Unknown (0%) |
| Alcohol use | Yes (3%); No (97%); Unknown (0%) |
| Smoking status | Yes (10%); No (90%); Unknown (0%) |
| Beta Blocker | Yes (25%); No (75%); Unknown (0%) |
| Calcium Channel Blocker | Yes (18%); No (82%); Unknown (0%) |
| Digoxin | Yes (10%); No (90%); Unknown (0%) |
| Diuretics | Yes (43%); No (57%); Unknown (0%) |

* After recoding 0's to missing values
